# SARS-CoV-2 in wastewater settled solids is associated with COVID-19 cases in a large urban sewershed

**DOI:** 10.1101/2020.09.14.20194472

**Authors:** Katherine E. Graham, Stephanie K. Loeb, Marlene K. Wolfe, David Catoe, Nasa Sinnott-Armstrong, Sooyeol Kim, Kevan M. Yamahara, Lauren M. Sassoubre, Lorelay M. Mendoza, Laura Roldan-Hernandez, Linlin Li, Krista R. Wigginton, Alexandria B. Boehm

## Abstract

Wastewater-based epidemiology (WBE) may be useful for informing public health response to viral diseases like COVID-19 caused by SARS-CoV-2. We quantified SARS-CoV-2 RNA in wastewater influent and primary settled solids in two wastewater treatment plants to inform the pre-analytical and analytical approaches, and to assess whether influent or solids harbored more viral targets. The primary settled solids samples resulted in higher SARS-CoV-2 detection frequencies than the corresponding influent samples. Likewise, SARS-CoV-2 RNA was more readily detected in solids using one-step digital droplet (dd)RT-PCR than with two-step RT-QPCR and two-step ddRT-PCR, likely owing to reduced inhibition with the one-step ddRT-PCR assay. We subsequently analyzed a longitudinal time series of 89 settled solids samples from a single plant for SARS-CoV-2 RNA as well as coronavirus recovery (bovine coronavirus) and fecal strength (pepper mild mottle virus, PMMoV) controls. SARS-CoV-2 RNA targets N1 and N2 concentrations correlate positively and significantly with COVID-19 clinical confirmed case counts in the sewershed. Together, the results demonstrate that measuring SARS-CoV-2 RNA concentrations in settled solids may be a more sensitive approach than measuring SARs-CoV-2 in influent.

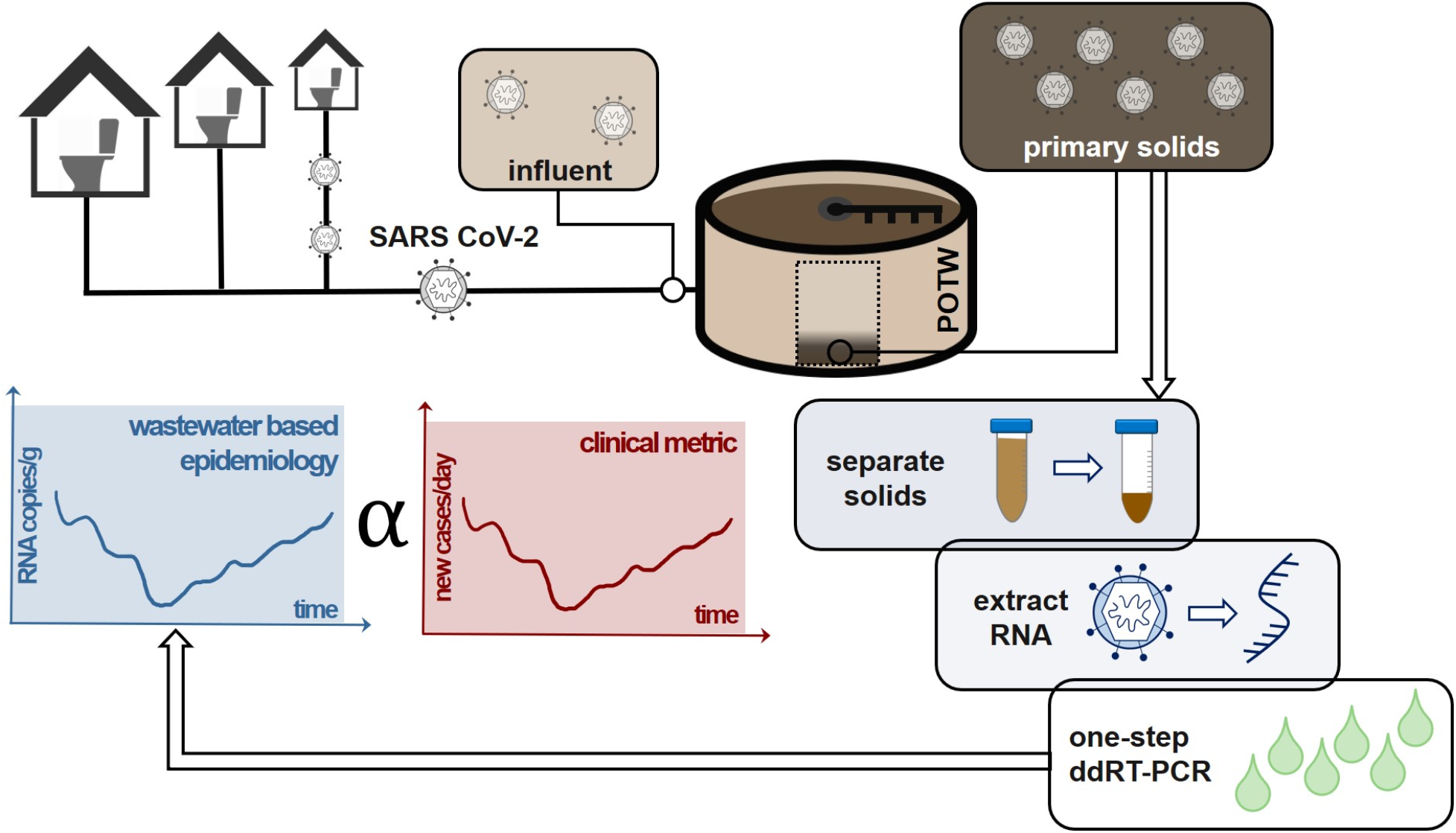

## Introduction

Municipal wastewater is a composite biological sample of an entire community with each member of the community inputting biological specimens to the wastewater every day. It is therefore no surprise that wastewater has been tapped as an epidemiological tool to gauge aspects of public health, such as narcotics usage ^1,2^, the reemergence of poliovirus ^3,4^, and infection rates of viral ^5,6^ and bacterial^7,8^ diseases. COVID-19 has accelerated the interest in wastewater-based epidemiology (WBE) due to the fact that SARS-CoV-2 genes are detected in the feces of many infected individuals^9–11^. More established epidemiological tools used to track cases in a community have been hindered during the COVID-19 pandemic by diagnostic kit shortages^12^, asymptomatic or mild cases that do not encounter the medical system or delay seeking medical attention^13^, and the lag times between testing and reporting^14^. As a result, public health officials and administrators have had to make critical decisions about opening or closing communities with limited surveillance data. Scientists, engineers, public officials, as well as the general public, are optimistic that WBE could provide additional data on COVID-19 infections in a community. In fact, the United States Center for Disease Control has established the National Wastewater Surveillance System as a framework for using WBE to inform the response to the COVID-19 pandemic ^15^.

Recent published studies have reported SARS-CoV-2 detection and quantification in sewage^16–18^. Based on these reports, numerous entities/organizations across the globe and across scales are moving to implement WBE. It remains to be seen how the data generated from wastewater surveillance should be interpreted or will ultimately be used to make public health decisions. Potential uses include informing on the presence or absence of COVID-19 in a community, similar to polio surveillance ^4^; tracking trends over time to project infection trajectory in the coming days^17^; or even using the SARS-CoV-2 concentrations in wastewater to estimate prevalence in a community^19^. The latter application requires a clear understanding of fecal shedding dynamics over the course of the illness, which is not yet established.

Many early studies on SARS-CoV-2 in wastewater have focused on wastewater influent. When detected in influent, viral particles are first concentrated with either a filtration method^18^ or an organic flocculation method^20^ and then the SARS-CoV-2 genome is targeted with a PCR-based method. In an earlier study, enveloped viruses mouse coronavirus murine hepatitis virus MHV and bacteriophage Phi6 partition to a greater extent to wastewater solids in wastewater influent than nonenveloped bacteriophages MS2 and T3^21^. The partition coefficients from that study were 1500, 1200, and 270 ml/g for MHV, Phi6, and MS2. These results suggest that wastewater solids may contain coronaviruses at concentrations 1000 times those found in influent, on a per mass basis, and that monitoring solids could lead to more sensitive detection of SARS-CoV-2^22^. Indeed, human coronaviruses HKU1 and 229E were previously detected in residual biosolids with metagenomic sequencing^23^.

In this study, we compared SARS-CoV-2 concentrations recovered from paired wastewater influent and primary settled solids collected at two different wastewater treatment plants collected on 5-7 days during a rising limb of the outbreak using different analytical methods. Subsequently, we applied refined methods to near daily samples of primary settled solids at a wastewater treatment plant over 89 days to investigate how SARS-CoV-2 concentrations in solids tracked COVID-19 cases in the sewershed. In addition to quantifying two SARS-CoV-2 RNA targets (N1 and N2), we quantified coronavirus recovery and wastewater fecal strength in every sample using bovine coronavirus (BCoV) and pepper mild mottled virus (PMMoV), respectively. Although the work focuses on a single pandemic virus, the results will be relevant for a wide suite of viral targets that have an affinity for solids.

## Methods and Materials

### Sample collection and storage

Influent and primary settled solids were collected from two publicly owned treatment works (POTWs), hereafter referred to as POTW A (Palo Alto Regional Water Quality Control Plant) and POTW B (San José-Santa Clara Regional Wastewater Facility), both located in Santa Clara County, California, USA. POTW A and B serve populations of 0.2 million and 1.5 million, respectively with permitted flows of 39 and 167 million gallons per day. POTW B adds FeCl_3_ to its waste stream (10 mg/l) prior to the headworks for odor control. The residence time of sludge in the primary clarifier is estimated to be between 2-14 hours for POTW A and 1-2 hours for POTW B.

From both plants, 50 ml of influent was collected at the headworks as 24-hour flow-weighted composite samples and 50 ml of solids samples were collected from the primary settling tank. Grab solids samples were collected from POTW A and 24-hour composite samples from POTW B. Samples were collected in 10% HCl acid-washed plastic containers. Storage conditions are described in the Supporting Information (SI).

Paired influent and solids samples were collected at both POTWs on the following dates: 22 March, 25 March, 29 March, 1 April, and 15 April 2020. Additional dates were included for POTW B (23 March and 30 March) (Table 1). Dates were chosen to span a high prevalence period in the initial phase of the pandemic. These samples are hereafter referred to as “method evaluation samples”.

**Table 1.** Inventory of samples used in this study. Unless specified in the column name, samples were processed using RT-QPCR and ddRT-PCR for N1 and N2, and RT-QPCR for PMMoV and BCoV. Unless otherwise specified, solids samples were extracted using the powerfecal kit.

| Date | Influent<br>POTW A<br>(Palo Alto) | Solids<br>POTW A | Influent<br>POTW B<br>(San Jose) | Solids<br>POTW<br>B | Solids<br>POTW B<br>(RNeasy) | Solids<br>POTW B 2-step dd-RT- PCR for N1/N2 |
| --- | --- | --- | --- | --- | --- | --- |
| 3.22.20 | X | X | X | X | X | X |
| 3.23.20 |  |  | X | X |  |  |
| 3.25.20 | X | X | X | X | X | X |
| 3.29.20 | X | X | X | X | X | X |
| 3.30.20 |  |  | X | X |  |  |
| 4.1.20 | X | X | X | X | X | X |
| 4.15.20 | X | X | X | X | X | X |

A longitudinal collection of primary settled solids was obtained from POTW B. Eighty-nine samples were collected daily from March 16 to May 31, and three times a week from June 2 to July 12, using the same collection techniques as the method evaluation samples. Hereafter, these are referred to as “longitudinal samples”.

### Influent pre-analytical processing

Immediately prior to analysis, influent samples were transferred to 4°C until thawed (range: 12-48 hours). Between 43 and 45 mL of influent was centrifuged at 24,000xg for 15 minutes at 4°C to pellet solids. 40 to 42 mL of the resultant clarified supernatant was decanted and viruses were concentrated from the supernatant using a PEG precipitation method^21^. This method was selected after preliminary head-to-head testing of PEG precipitation and centrifugal ultrafiltration to concentrate viruses from influent yielded comparable recovery results for N1 and N2 (Figure S1), and because we anticipated there would be fewer pandemic-related supply chain issues with the PEG method. MHV strain A59 and PMMoV were used as an exogenous and endogenous recovery controls, respectively. PMMoV also served as a fecal-strength control. An attenuated vaccine strain of bovine coronavirus (BCoV) was used as a nucleic-acid extraction positive control; it was spiked into the viral concentrate before it was subjected to nucleic-acid extraction using the Qiagen AllPrep PowerViral DNA/RNA kit and further purification using Zymo OneStep PCR Inhibitor Removal Columns (Zymo Research, Irvine, CA). Further details are in the SI.

### Solids pre-analytical processing

Immediately prior to analysis, solid samples were transferred to 4°C until thawed (range: 12-48 hours). Thawed solids samples (35-53 ml) were centrifuged at 24,000xg at 4°C for 30 minutes, and the supernatant was decanted. Percent solids was measured for concentrated solid samples by placing sample aliquots in pre-weighed aluminum weigh dishes and weighing the sample before and after drying at 105°C for 24 hours.

The method evaluation solids samples were divided to test two different extraction processes: for the first, 0.2-0.4 g of sample was aliquoted for extraction with the Qiagen AllPrep PowerFecal DNA/RNA kit (“powerfecal” kit), and for the second, 1.8-2.8 g of sample was aliquoted for extraction with the RNeasy PowerSoil Total RNA Kit (“RNeasy” kit). Aliquots were spiked with BCoV as an extraction recovery control. Two biological replicates were completed per sample (2 powerfecal and 2 RNeasy extractions per POTW per time point). Nucleic-acids were further purified using the Zymo OneStep PCR Inhibitor Removal Columns (Zymo Research, Irvine, CA). Further details are in the SI.

For the longitudinal solids samples, RNA was extracted using the RNeasy kit and purified with the Zymo inhibitor removal kit using the same protocol described above with two modifications ^24^; (1) after step 9 in the kit’s instructions, samples were stored overnight at -20°C, and (2) positive pressure was used per manufacturer’s instructions to aid flow through columns. 7 of the 89 samples were extracted in duplicate and extraction blanks were run with each sample batch. Samples were randomized and blinded to laboratory technicians prior to analysis.

### RT-QPCR

For method evaluation samples, PMMoV, MHV, BCoV, SARS-CoV-2 N1 and N2 were quantified using two-step RT-QPCR. Preliminary experiments suggested that the RT step was inhibited, so dilutions of the extract were used as template for the RT step (see SI for details). For influent, undiluted and 1:10 diluted RNA was used as template; for solids, undiluted, 1:10, and 1:50 diluted RNA were used as template. Each template was run in triplicate, and triplicate reactions were pooled prior to QPCR.

Resultant cDNA was used as template in QPCR assays targeting PMMoV, MHV, BCoV, SARS-CoV-2 N1 and N2 (primers and probes in Table S1) using an ABI Step-One Plus Instrument (Thermo Fisher Scientific, Waltham, MA). All templates were run in duplicate. Standard curves were run in duplicate on each plate using cDNA at concentrations from 3 to 3 × 10^5^ copies / reaction; duplicate NTCs were included on each plate. Detailed methods for generation of cDNA standards are in the SI. Concentrations per reaction were converted to copies per volume of influent or per g dry solids weight using dimensional analysis. Samples were considered “below the limit of quantification” (BLOQ) if the Cq value of the sample was higher than the Cq value of the lowest cDNA standard, but less than 40, or if the targets in both technical replicates did not amplify within 40 cycles. Concentrations of unknown samples were calculated using plate-specific standard curves. Additional details are in the SI^25^.

### ddPCR

RNA from the method evaluation samples was used as template in one-step digital droplet (dd)RT-PCR for N1 and N2 SARS-CoV-2 targets using BioRad SARS-CoV-2 Droplet Digital PCR kits (cat.no.: 12013743; Hercules, CA) and a BioRad QX200 AutoDG Droplet Digital PCR system. Technical duplicates were run (2 wells); wells were merged for data analysis. In order to test for inhibition, each template was run at two dilutions: undiluted and 1:10 dilution. A positive control consisting of SARS-CoV-2 RNA isolated from a nasopharynx swab of a high-titer patient from Stanford Hospital and NTCs were run on each plate in duplicate. QuantaSoft and QuantaSoft Analysis Pro (BioRad) were used to manually threshold and export the data. The required number of droplets for a sample with merged duplicate wells was at least 10,000. In order for a sample to be scored as positive, 3 or more positive droplets were required in the merged wells. Concentrations per reaction were converted to copies per volume of influent or per g dry weight using dimensional analysis.

A two-step ddRT-PCR assay for N1 was also trialed using a subset of the method evaluation samples. RNA was stored at -80°c for up to 5 days between running the one-step and two-step assays, to minimize the potential for RNA degradation during storage (see SI).

For the longitudinal samples, one-step ddRT-PCR duplex assay for N1 and N2 and a duplex assay for PMMoV and BCoV^26^ were used. N1 and N2 were quantified in undiluted and 1:10 diluted template; BCoV and PMMoV were quantified in 1:10 and 1:1000 diluted template. The N1/N2 duplex assay was conducted in triplicate wells and the BCoV/PMMoV duplex assay was conducted in duplicate wells. The replicate wells were merged for data analysis. Positive N1/N2 and negative controls, and threshold setting for the longitudinal samples were conducted as described for the method evaluation samples. Positive controls for BCoV and PMMoV were a direct extraction of reconstituted BCoV vaccine diluted to 10^6^ cp/mL and for PMMoV a synthetic DNA ultramer. In addition, a pooled matrix control was run on every N1/N2 duplex assay plate. This was created by adding equal-volume aliquots of each sample on the plate to one tube and spiking this pooled matrix with clinical SARS-Cov-2 RNA. Additional details of dd-RTPCR^27^ can be found in the SI and Table S7.

### COVID-19 epidemiology data

The number of new clinically confirmed COVID-19 cases within the sewershed of POTW B were obtained using geo-referenced case data and POTW-provided shape file of their service area in ARC-GIS. The date stamp is that of specimen collection. As data were handled and provided by the county, no IRB approval was needed.

### Statistics

Statistics were computed using Microsoft Excel and Rstudio (version 1.1.463). Paired and unpaired t-tests were used to compare groups after confirming data were normally distributed. For calculating recoveries, and making numerical comparisons across samples using the high copy number targets measured by RT-QPCR (PMMoV, MHV, and BCoV), the results from reactions that yielded the highest measured concentration were used to balance loss of signal against effects of inhibitory substances. For ddRT-PCR data, “total error” from merged wells is reported in the form of standard deviations. Pearson correlation coefficients assessed association between different measurements among samples.

We used a resampling-based strategy to evaluate whether the observed measurements N1 and N2 with and without PMMoV normalization were predictive of clinically confirmed cases. We applied first differencing to account for autocorrelation, resulting in the following regression specification (using N1 as the example): Cases_t_ − Cases_t-k_ ~ N1_t_ – N1_t-k_ where the subscript t indicates the day of measurement and k is the number of days used for differencing, in this case 7; k = 14 was also used as a sensitivity test. If a wastewater data point was missing at day t-k, then the value for the closest date was imputed. In order to account for the technical variability of the measurements, the observed top and bottom confidence bounds were used to resample wastewater concentrations uniformly. For samples below the detection limit, we used uniform sampling between 0 and the lower detection limit (estimated at 40 copies/g dry weight). In addition to raw case counts, 7-d smoothed case data were also used to account for the day-to-day variation in case data. A downsampling analyses was conducted to investigate the frequency of sample collection needed to observe associations between case counts and wastewater data. We downsampling to fortnightly, weekly, and twice-per-week frequencies using a random observation within each appropriate period and examined as described above except k for biweekly was are 2 or 4 half-weeks, and for weekly and fortnightly the k was 1 or 2 (weeks/fortnights). Empirical p-values were determined with 1,000 resamplings were reported.

## Results and Discussion

### Analytical controls and assay performance

NTCs and extraction blanks were negative for all targets in all analytical methods. QPCR assay efficiencies ranged from 78-105% and standard curve R^2^ ranged from 0.98 to 0.99 (Table S2; Figure S2).

### Method Evaluation: Comparison of RT-QPCR vs ddPCR assays on influent and settled solids

With RT-QPCR, N1 and N2 were not detected in any influent sample, even when RNA template was diluted 1:10 and cDNA was diluted 1:10, except for a single influent from POTW B (Table S3) that resulted in detection below the limit of quantification. When one-step ddRT-PCR was applied to the same undiluted and 1:10 diluted RNA templates, one sample tested positive for N1 in POTW A, three tested positive for N1 at POTW B, and one tested positive for N2 in POTW B (Table S4). In all but one of the samples that resulted in positive measurements, the undiluted template yielded a non-detect, suggesting inhibition of the reactions in the undiluted template.

For solids samples extracted with powerfecal kits and analyzed with RT-QPCR, SARS-CoV-2 targets were detected in at least one biological replicate from 1 of 5 samples from POTW A and 4 of 7 samples from POTW B (Table S3). Detection was inconsistent across biological and technical replicates, and there was no case where both biological or technical replicates were positive. When analyzed with ddRT-PCR, the SARS-CoV-2 targets were detected and quantified in 0 out of 5 POTW-A solids samples and 7 out of 7 POTW B samples (Table S4). ddRT-PCR results were more consistent across biological replicates than RT-QPCR (Figure 1; Table S4).

**Figure 1.**
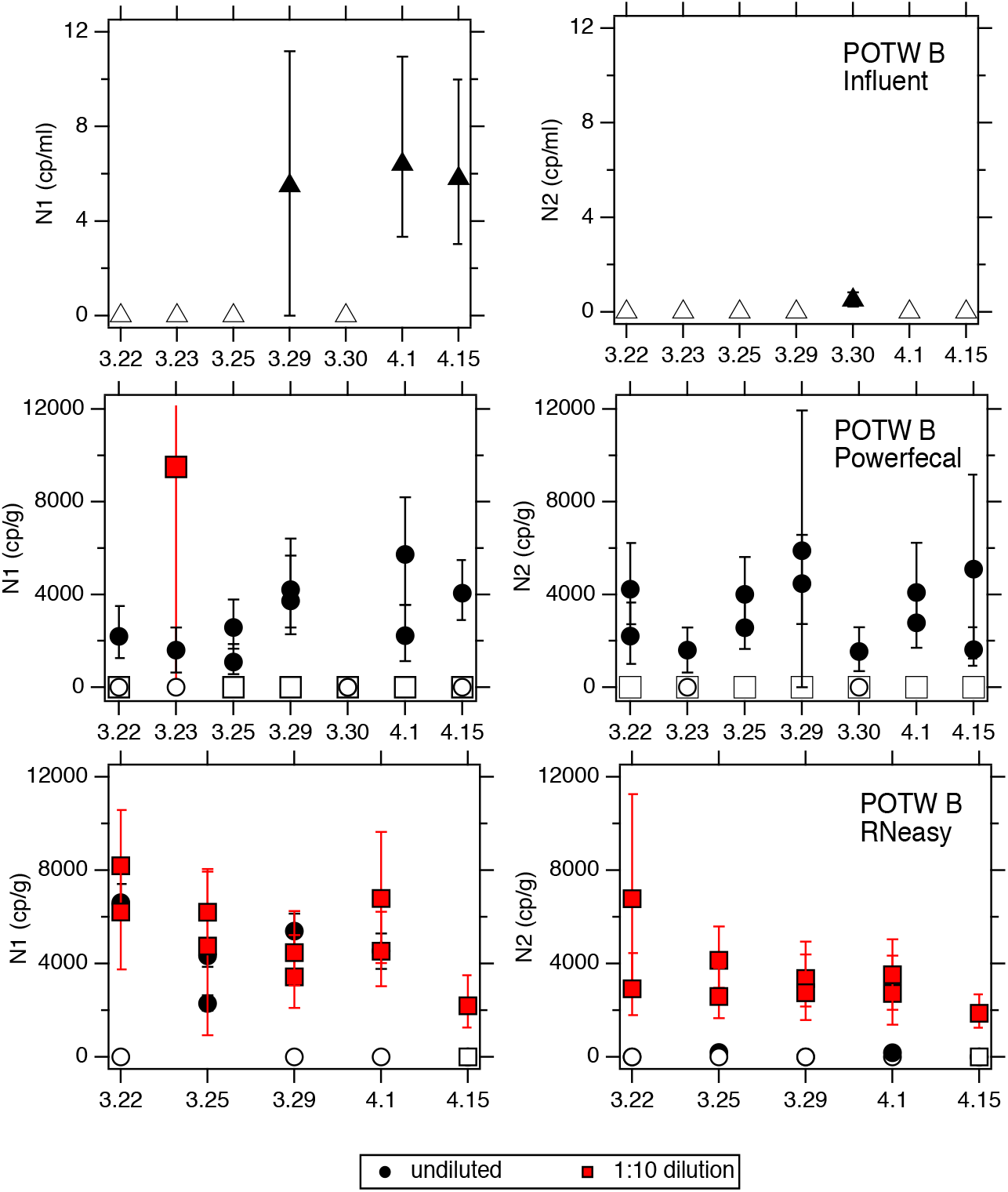
N1 and N2 measured at POTW B using ddRT-PCR. Top row: Concentrations in influent in units of copies (cp) per ml. Middle row: Concentrations in solids extracted using the powerfecal kit in units of cp per g dry weight. Bottom row: Concentrations in solids extracted using the RNeasy kit in units of cp per g dry weight. Open symbols indicate those where no target was detected (less than 3 positive droplets). Error bars represent standard deviations as represented by the “total error” which includes Poisson error as well as differences among merged wells. Results for undiluted and 1:10 diluted template are provided for the solids; results for influent are for 1:10 diluted template for N1 and undiluted template for N2 – results for dilutions not shown are all non-detects.

The improved performance of ddRT-PCR compared to RT-QPCR for both influent and solids samples was unlikely due to differences in template amounts. The virtual influent volumes in RT-QPCR and ddRT-PCR assays were 2 mL and 8.4 mL, respectively (when template not diluted); and for solids, the virtual wet mass of solids assays in the RT-QPCR and ddRT-PCR reactions were 20 mg and 50 mg, respectively (when template not diluted); thus theoretical lower detection limits are similar across analytical approaches (Table S5). There are several possible explanations for the low and inconsistent detection of SARS-CoV-2 targets in influent and solids with RT-QPCR, including assay inhibition, and the absence or low occurrence of viral RNA in the samples. We studied RT-QPCR inhibition in more depth using endogenous PMMoV and spiked MHV and BCoV (Figure S3). The results demonstrate inhibition for the RT-QPCR assays of endogenous and exogenous recovery targets, despite efforts to alleviate it using the Zymo OneStep Inhibitor Removal columns and dilution of template. For influent, PMMoV and MHV concentrations were generally highest when RNA template was diluted 1:10 prior to RT, compared to results from undiluted and 1:10 diluted cDNA generated from undiluted RNA template (Figure S3). This suggests the RT step, and not the PCR step, was most inhibited for the PMMoV and MHV RT-QPCR assays. For solids, extract dilutions of 1:50 appeared to alleviate inhibition for RNA obtained from POTW A (Figure S3; Table S6). For solids from POTW B, however, inhibition may not have not alleviated even after a 1:50 dilution.

Based on these results comparing the performance of RT-qPCR and ddRT-PCR, and the improved sensitivity of the ddRT-PCR, we selected the ddRT-PCR methods for further SARS-CoV-2 analysis.

### Method Evaluation: Comparison of RNeasy and powerfecal extraction kits and 1-step vs. 2-step ddPCR for settled solids

We used the SARS-CoV-2 ddRT-PCR assays and the POTW B solids to compare powerfecal and Rneasy solids extraction kits. The virtual wet mass of solids assayed in the duplicate merged wells was approximately 50 mg (powerfecal) and 250 mg (RNeasy) when the templates were not diluted (theoretical detection limits in Table S5). Samples treated with the powerfecal kit were more commonly positive for SARS-CoV-2 targets in undiluted template than in 1:10 diluted template (Figure 1). By contrast, samples treated with the RNeasy kit were positive for SARS-CoV-2 at both dilutions, but detection was more consistent with the 1:10 diluted templates (Figure 1). The SARS-CoV-2 target concentrations were similar across the biological replicates and the null hypothesis that paired replicates have the same concentration was not rejected (p>0.05 for all 4 comparisons (2 targets × 2 RNA extraction kits), paired t-test). The concentrations of N1 and N2 measured in the same sample / biological replicate were not different between the two kits (paired t-test, p>0.05). Although the powerfecal kit samples less material than the RNeasy kit, these results suggest that some of the RNeasy kit extracts require a 1:10 dilution, thus negating the advantages of the extra sample mass.

Whereas the N1 target was detected in every solids sample from POTW B when the 1-step ddRT-PCR assay was employed, it was not detected in any of the RNeasy extracts when a 2-step ddRT-PCR assay was conducted.

### Method Evaluation: Comparison of SARS-CoV-2 RNA concentrations measured in influent and primary sludge

We compared SARS-CoV-2 measurements made in influent and solids samples collected on the same days at POTW B (Figure 1). Solids results obtained using the powerfecal kits are used for the comparisons. At POTW B, the N1 target was detected in 3 out of 7 influent samples and 6 out of 7 solids samples and the N2 target was detected in 1 out of 7 influent samples and 7 out of 7 solids samples. The N1 target was quantified in both influent and primary solids on 3 dates (3/29, 4/1, 4/15) and the N2 target was quantified in both influent and primary solids on 1 date (3/30). The ratio of N1 solids concentrations (cp/g; mean of biological replicates) to influent concentrations (cp/mL) on the 3 dates was 720, 620, and 350 ml/g respectively. For the N2 target, the ratio was 3100 ml/g. These results suggest that on a per mass basis, these targets are present at between ~100 and ~1000 higher concentrations in solids.

### Method Evaluation: Recovery of viral targets through preanalytical processing

Recovery measurements help characterize preanalytical method consistency across samples. For the influent method, we measured the recoveries of endogenous PMMoV and spiked MHV through the PEG concentration method (Figure S4). Median PMMoV recovery was 21% (range: 8% to 30%) and median MHV recovery was 7% (range: 2% to 16%). MHV recovery was lower than that for PMMoV by, on average, 13% (paired t-test, p<0.05). We used BoCoV to assess recovery specifically through nucleic-acid extraction and purification, and through inhibitor removal steps. Recovery of BCoV through these steps ranged from 3% to 48% (median= 26%). The PEG concentration step and nucleic acid extraction/purifications step are conducted in series. Assuming that the BCoV extraction recoveries apply to all viral targets, our results suggest that viral recoveries are between 0.1-7% (median 1%, based on MHV) or 1-8% (median 7%, based on PMMoV) of actual influent concentrations.

There were no significant differences in recoveries of PMMoV, MHV, and BCoV in influent between the two POTWs (t-test, p>0.05), or in samples that were positive or negative for N1 or N2 (t-test, p>0.05). In fact, the influent sample with one of the lowest MHV and BCoV recoveries (POTW B on 3/29/20) was one of the few influent samples positive for N1.

PMMoV may serve not only as a process control, but also a means of assessing the “fecal strength” of sewage. PMMoV measured in the viral concentrate varied between 7.7 × 10^2^ to 7.0 × 10^3^ copies/ml of influent across plants (Figure S4). There was no significant difference in PMMoV influent concentrations between plants (p>0.05).

### Method Evaluation: Endogenous and spiked virus recoveries in solids from method evaluation study

There was no viral concentration steps in the solid methods and BCoV served as an extraction recovery control. BCoV recovery varied from 0% to 33% (median = 16%) using the powerfecal kit and 0% to 48% (median = 22%) using the RNeasy kit (Figures S5 and S6). The 0% recovery values were obtained in POTW B solids collected on 15 April and treated with the RNeasy kit and POTW B solids collected on 25 March and treated with the powerfecal kit. Interestingly, PMMoV and SARS-CoV-2 targets were measured in the same extracts with 0% BCoV recovery at concentrations not largely different from other dates. The measured BCoV recoveries were not different across plants or across extraction kits (t-test, p>0.05). It is possible, however, that the BCoV recoveries in the POTW B solids are underestimates because we are not certain that the RT-QPCR assay conducted at the highest extract dilution was free of inhibition.

PMMoV in solids from POTW A and POTW B varied between 5×10^4^ and 7×10^7^ copies/g dry weight (median 2 × 10^6^ copies/g) when extracted with powerfecal kit (Figure S6). Only samples from POTW B were measured with the RNeasy extraction kit and the resulting concentrations ranged from 5×10^5^ to 3×10^6^ copies/g dry weight (median 1 × 10^6^ copies/g; Figure S5). These concentrations are twice as high, on average, than those measured using the powerfecal templates (paired t-test, p<0.05). The PMMoV concentrations may be underestimates due to unresolved assay inhibition. Concentrations of PMMoV were not different across biological replicates regardless of which extraction kit was used (paired t-test, p>0.05). PMMoV concentrations measured in solids at POTW A were not different from those measured at POTW B (t-test, p>0.05, data from powerfecal extracts).

N1 and N2 concentrations in solids are not correlated to BCoV recovery or PMMoV concentrations regardless of kit used (p>0.05) (Figures S5 and S6).

### Longitudinal data from POTW B

The method performance experiments demonstrated that the primary solids method resulted in more positive and consistent SARS-CoV-2 signals than the influent method. Furthermore, we found that the 1-step ddRTPCR assay performed on the solids extracts resulted in more positive results than the 2-step RT-qPCR and 2-step ddRTPCR assays. We therefore focused our longitudinal analysis on primary settled solids samples from POTW B using the RNeasy kit and ddRT-PCR for measuring all targets (Figure 2).

**Figure 2.**
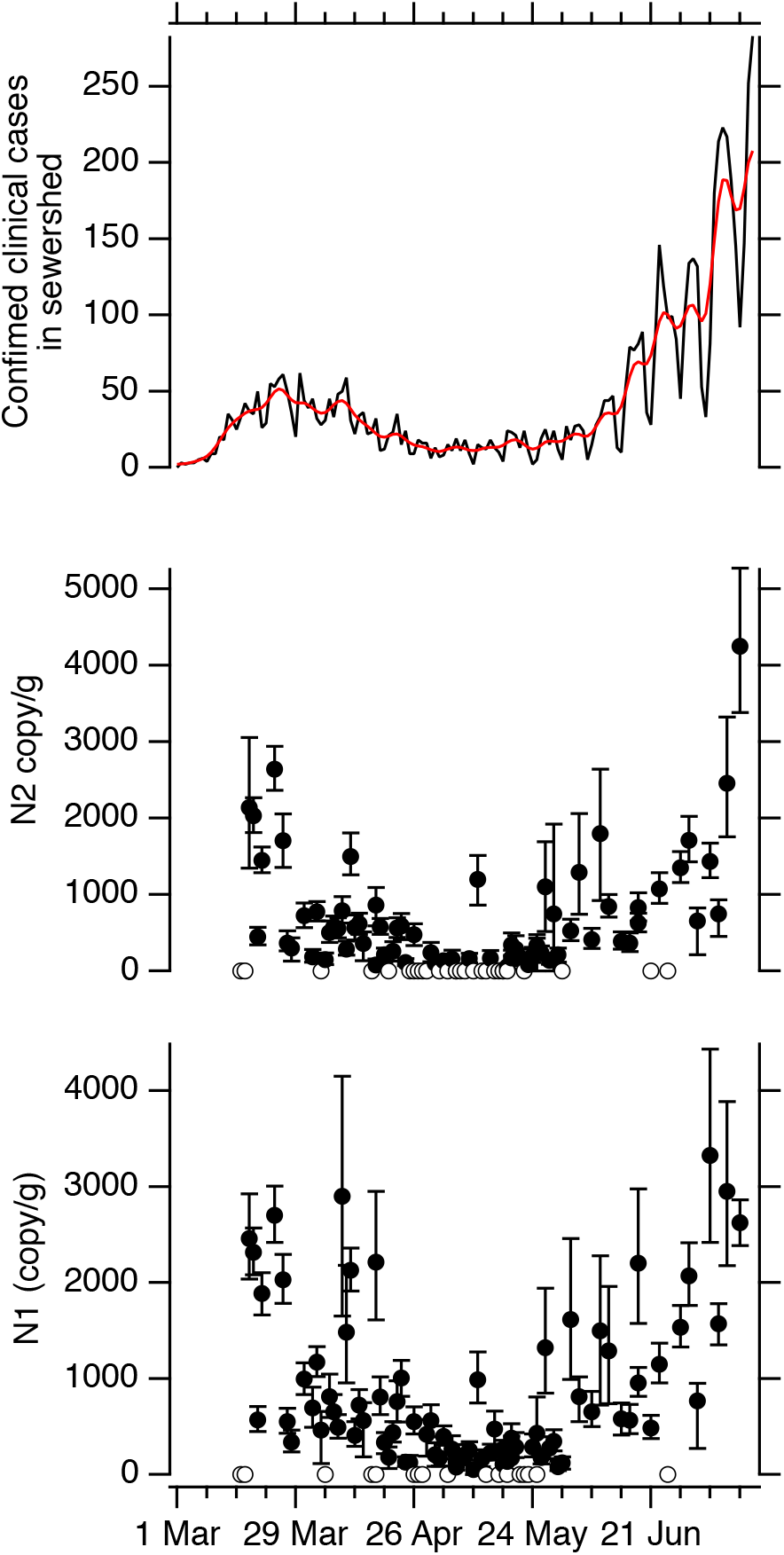
Top panel: Confirmed new cases of COVID-19 in the sewershed of POTW B (black) and 7-d smoothed new cases (red). Middle and bottom panels: Concentrations of N1 and N2 measured in solids (copy per g dry weight). Error bars are standard deviations as total errors from the ddPCR machine. Open symbols are non-detects plotted at 0. The theoretical lowest concentration measurable is ~40 copies/g dry weight.

Concentrations of N1 and N2 in each sample were derived from the reactions that used undiluted RNA template unless those that used 1:10 diluted template produced a concentration higher than the upper standard deviation for the undiluted template (10/96 and 6/96 samples required 1:10 dilution for N1 and N2, respectively).

N1 and N2 were detected in 79 and 68 of the 96 samples. When detected, average N1 and N2 values were 870 cp/g and 730 cp/g with maximums of 3330 cp/g and 4250 cp/g, respectively. Replicate extractions generally yielded concentrations that were similar.

Median PMMoV was 2×10^7^ cp/g (interquartile range 1 ×10^7^ cp/g to 4 ×10^7^ cp/g) (Figure S7) suggesting fairly constant fecal strength of the solids. We normalized measured concentrations of N1 and N2 to obtain fecal strength normalized target concentrations to include in the statistical analyses. The recovery of BcoV was lower in the longitudinal samples than it was in the method evaluation samples, with a median recovery was 3% (interquartile range 1 to 5%) (Figure SA). We omitted the N1 and N2 data from our dataset for the one sample where 0% BCoV recovery was measured (16 March). Recovery was higher than 10% on 8 days; N1 and N2 concentrations were not significantly different on those days compared to the others (p>0.05).

The concentrations of N1 and N2 tracked the reporting of new cases in the sewershed (based on specimen date) when accounting for autocorrelation and technical errors associated with the wastewater measurements (using raw case counts, 0.024 cases / copies /g for both N1 and N2, p = 0 and 0.005, respectively). Results were not sensitive to choice of k, smoothing of case data, or normalization of N1 and N2 by PMMoV to account for changing sample-to-sample differences in fecal strength or recovery (Table S8). Downsampling wastewater data collection and analysis frequency to down sampling to twice-per-week yields significant associations between case counts and wastewater concentrations; however, downsampled to fortnightly or once per week, associations are no longer significant (Table S8).

## Discussion

Our results suggest that testing wastewater solids for SARS-CoV-2 may be more sensitive than testing influent. We detected co-occurring SARS-CoV-2 N1 and N2 targets in nearly every solid sample tested at POTW B, yet both targets were never co-detected in influent samples. The lack of detection of SARS-CoV-2 targets at POTW A in both influent and solids may be a result of low prevalence of COVID-19 in the relatively small sewershed. The measured ratios of N1 and N2 concentrations in solids and influent were between 350 and 3100 ml/g; these values are likely low estimates as the influent N1 and N2 concentrations were below our detection limit in most of the samples from many days when N1 and N2 were detected in solids. Depending on the pre-analytical method used in our study (i.e., extraction kit, template dilution factor, etc.), there was between 20 and 1100 times more influent than dry solids on a per mass basis in the PCR reactions. Given that solids have, at minimum, between 350 and 3100 higher concentrations, the number of target copies could be much higher in the PCR reactions for solids compared to influent. The increased sensitivity in detecting SARS-CoV-2 in solids will be particularly important for screening communities at the sewershed scale for outbreaks occurring within communities with low levels of COVID-19 prevalence. Other viruses have a high affinity for wastewater solids including adenovirus, rotavirus, and various enteroviruses^28–31^, so solids are likely to be useful for WBE applied to other viral diseases. We also note that solids analysis avoids the preconcentration step necessary with influent analysis.

Using longitudinal samples from POTW B, we identified a positive association between N1 and N2 in solids and the number of new COVID-19 infections. A similar result was reported by Peccia et al.^22^ for a POTW on the east coast of the US. When N1 and N2 are normalized by fecal strength, as determined by PMMoV, associations were similar. Whether the N1 and N2 signals lead the new case data requires further analysis complicated by the autocorrelation of the data. Evaluating multiple POTWs will improve robustness of such estimates. The positive associations suggest that SARS-CoV-2 RNA in solids can be used to confirm trends in infection prevalence; and therefore, augment case data to inform a response to the COVID-19 pandemic.

Ideally, wastewater surveillance could take place with high resolutions sample collection (e.g., daily); however, this may not be possible for many utilities/communities due to limited staffing, supplies, and resources. Our down-sampling analysis suggests sampling solids twice per week would be frequent enough to identify the global trends in the clinical case data. It is important to note that case data itself are imperfect as testing availability, population testing bias, and time to testing result can vary substantially. For example, during the initial phase of the pandemic (between mid-March to mid-April), clinical testing was not widely available to people residing in the sewershed; however, sentinel surveillance efforts suggest that during this time that there was transmission in the community^32^. While prevalence in the sewershed during this period is unknown, given limited testing, interestingly, solids concentration of N1 and N2 are just as high during the initial infection peak as during the second infection peak and may represent an additional, less biased source of monitoring data.

Whereas Peccia et al.^22^ recently reported correlations between N1 and N1 in sludge and new COVID-19 cases during a period when infections were rising, we report correlations across both rising and falling periods of the epidemiological curve. Given that individuals likely shed SARS-CoV-2 for weeks^33,34^, it is somewhat surprising that SARS-CoV-2 RNA concentrations correlate with the number of new COVID-19 infections. This result may yield insight into the time course and magnitude of fecal shedding and is consistent with substantially higher shedding rates during the initial phase of the infection^35^. More fecal shedding data are needed, especially for pre-symptomatic individuals to fully understand if SARS-CoV-2 RNA concentrations in solids can be linked quantitatively to prevalence of infection in the sewershed.

The RNA extraction, concentration, and purification procedures used in this study did not eliminate RT and PCR inhibition, despite the use of clean-up kits. Dilution of templates was needed, at times, for both influent and solids to obtain signals, even using ddRT-PCR which is known to decrease the effects of inhibition in some cases^36,37^. Wastewater is a diverse, complex matrix that differs chemically and biologically between POTWs. Presence of metals, organics, and other material can impede the reverse transcription and PCR ^38^. The solids from POTW B appeared to be more highly inhibited than those from POTW A, likely because they contain Fe-containing compounds as a result of their FeCl_3_ addition, a known PCR inhibitor^39^. These results highlight the importance of testing for inhibition in wastewater matrices and also the need for more research on how to alleviate inhibition without diluting the extracts.

We used recovery controls in our analyses including an endogenous, non-enveloped virus (PMMoV), and two beta coronaviruses (MHV and BCoV). The recoveries we report are similar to those reported by others^40–43^. We did not correct concentrations reported in this study using measured recoveries as there is great uncertainty regarding whether a recovery surrogate mimics the behavior of the virus of interest – in this case SARS-CoV-2, even if it shares certain biological characteristics. It is impossible to know confidently that the spiked recovery target will interact with the components of the matrix in the same manner that an endogenous target interacts. In addition, it is not known whether the spiked recovery control mimics the state of the endogenous SARS-CoV-2 targets as it is unknown whether the SARS-CoV-2 virus exists as an intact virion in the sewage matrices, or if its lipid envelope is absent, or if its RNA is located outside of a lysed capsid. In light of all these complexities, we believe that recovery of a surrogate should be used to ensure that sample processing is not “out of the ordinary”, and can be used to identify problems during sample processing. Indeed, during the longitudinal sample processing we identified 1 sample where the BCoV recovery control failed, and removed that sample’s data from subsequent statistical analysis.

Surprisingly, we found that recovery for the longitudinal samples was reduced compared to that for the method evaluation study suggesting that the modifications we made to reduce processing time adversely affected recovery. An unanticipated benefit of the reduced recovery was reduced inhibition. During the methods evaluation study, the RNeasy-processed solid samples contained inhibitors such that N1 and N2 had to be measured in 1:10 dilutions of the template. During the longitudinal study, N1 and N2 could be measured in the undiluted template suggesting less inhibitors were co-extracted. This illustrates a tradeoff between inhibition and recovery. The reduced recovery observed for the longitudinal samples also explains why concentrations of N1 and N2 were higher by about a factor of ~4 at POTW B during method evaluation compared to samples from similar dates during the longitudinal study.

The amount of feces or urine in wastewater may vary with time based on sewershed populations, precipitation, changing waste streams, or human behaviors^44^. Such variation may affect targets relative to WBE and in this case, SARS-CoV-2 RNA. Normalization refers to the normalization of a WBE target by another wastewater target that is a measure of the amount of fecal material in the wastewater. Normalization is not a practice that has been extensively tested in environmental virology. However, it has been explored in the use of wastewater-based epidemiology for drug use ^44-46^. We tested PMMoV as a normalization factor as it is found extensively in wastewater globally and it originates in human feces^47^. PMMoV concentrations measured in the influent and solids of the plants we sampled did not vary substantially across samples or plants^47^ suggesting limited changes to the fecal content of the wastestream during the study. This may explain why normalizing N1 and N2 by PMMoV did not substantially change correlations with new COVID-19 cases in the sewershed.

## Implications and future work

Solids naturally concentrate SARS-CoV-2, making it a reliable target for WBE. However, there are situations where solids analysis may be impractical. For example, POTWs with no primary settling step or for sewage sampling from manholes within a sewershed, solids may be difficult to.

There are several avenues of research needed to further understand the limitations and utility of WBE for COVID-19. First, more information is needed about the decay of viral targets both in influent^48^ as well as within solids. This information is needed to better understand how infection in the sewershed affects concentrations in wastewater matrices. Second, the degree to which settled solids represent a natural composite sample owing to mixing processes in the settling tank need would be useful. While composite samples of influent are preferred over grab samples owing the changes in the influent stream with time ^49^, the same may not be true for settled solids eliminating the need for composite samplers. Importantly, additional information on duration and magnitude of shedding of viral RNA via feces is needed; as well as the extent to which viral RNA is encapsulated versus extravirual in feces and sewage is needed to predict its fate and transport in the wastestream.

## Data Availability

Data are available for the work by contacting authors.

## Supporting Information

The Supporting Information is available free of charge. Additional details of methods, ancillary tables (Tables S1 through S8) and figures (Figures S1-S8).

## Acknowledgments

This work was partially supported by NSF RAPID (CBET-2023057) grant to KW and AB. We thank B. Pinsky and M. Sahoo in Stanford Clinical Virology, as well as an anonymous patient, for providing our positive control SARS-CoV-2 RNA. N.S.-A. was funded by a Stanford Graduate Fellowship. This study was performed on the ancestral and unceded lands of the Muwekma Ohlone people. We pay our respects to them and their Elders, past and present, and are grateful for the opportunity to live and work here.

## Notes

### Competing Interest Statement

The authors have declared no competing interest.

### Author Declarations

No IRB approval needed for this work. No human subjects data.

